# Sex ratios and ‘missing women’ among Asian minority and migrant populations in Aotearoa/New Zealand: A retrospective cohort analysis

**DOI:** 10.1101/2021.03.11.21253424

**Authors:** Rachel Simon-Kumar, Janine Paynter, Annie Chiang, Nimisha Chabba

## Abstract

**Background:** Recent research from the UK, USA, Australia, and Canada point to male-favouring Sex Ratios at Birth (SRB) among their Asian minority populations, attributed to son preference and sex-selective abortion within these cultural groups. The present study conducts a similar investigation of SRBs among New Zealand’s Asian minority and migrant populations, who comprise 15% of the population.

**Methods:** The New Zealand historical census series between 1976-2013 was used to examine SRBs between ages 0-5 by ethnicity. A retrospective birth cohort in New Zealand was created using the Stats NZ Integrated Data Infrastructure from 2003-2018. A logistic regression was conducted and adjusted for selected variables of interest including visa group, parity, maternal age and deprivation. Finally, associations between family size, ethnicity and family gender composition were examined in a subset of this cohort (families with 2 or 3 children).

**Results:** There was no evidence of ‘missing women’ or gender bias as indicated by a deviation from the biological norm in New Zealand’s Asian population. However, Indian and Chinese families were significantly more likely to have a third child if their first two children were females compared to two male children.

**Conclusion:** The analyses did not reveal male-favouring SRBs or any conclusive evidence of sex-selective abortion among Indian and Chinese populations. Based on this data, we conclude that in comparison to other western countries, New Zealand’s Asian migrant populations presents as an anomaly. The larger family sizes for Indian and Chinese populations where the first two children were girls suggested potentially ‘soft’ practices of son preference.

**WHAT IS ALREADY KNOWN ON THIS SUBJECT:**

- There are discrepancies in Sex Ratios at Birth (SRB) among the Asian minority migrant populations – particularly Indian and Chinese populations –in countries like Canada, UK, USA. SRBs show a pronounced number of males over female children, suggesting a widespread practice of sex-selective abortions in these communities since the 1970s.
- These trends implicitly reflect social norms of gender bias through son preference, and daughter devaluation.

**WHAT IS BEING ADDED:**

- The present study did not find evidence of skewed SRBs that favour boys over girls among Asian ethnicities. The analyses however did find a tendency for Indian and Chinese families to have larger families especially when the first two children were girls.
- Overall, the findings suggest the absence of widespread practices of sex-selective abortion making New Zealand an anomaly relative to other migrant-receiving countries. However, there are still vestiges of son preference that are seen through decisions around family size and gender composition.

## BACKGROUND

Sex ratios at birth (SRB), calculated as Males:Females (M:F), are widely used as a measure of gender bias within a population.[1–3] Landmark work conducted some four decades ago in India and China highlighted imbalances in M:F birth ratios above the normative 105:100, an artefact of deliberate sex selection resulting in excess prenatal female mortality estimated at millions.[2, 4, 5] Widely referred to as the ‘missing women’ phenomenon, skewed M:F ratios are unequivocally accepted as reflecting son preference and undervaluation of girls and women; as Sen [5] noted, they “tell us, quietly, a terrible story of inequality and neglect” (p61).

Extensions to this scholarship more recently has focused on migrants of Asian heritage residing in ‘western’ societies. Burgeoning work in Canada,[6–14] the United Kingdom,[15–18] the United States of America,[19–24] Sweden,[25] Norway,[26] Australia,[27] and Spain,[28] has documented male bias in sex ratios largely among their Indian, Chinese and Korean migrant communities. Overall, analyses of sex ratios in these diverse country contexts estimate anywhere from 2,000 to 4,000 ‘missing women’ between the 1980s and 2000s.

Skewed M:F ratios among migrant Asian communities largely present among first-generation migrants, especially where the mother is overseas-born [8, 10, 13, 15, 27] although recent research in Canada shows continuation of these trends among second-generation South Asian women.[6] Studies point to imbalances in M:F ratios at higher parities (> 2) especially where the first two children are girls.[13, 15, 20, 22–24, 27, 29, 30] The practice is found among affluent, urban migrants, ruling out economic need as the exclusive driver of sex selection.[17, 18] Instead, residual cultural norms are pivotal in the drive for male children; traditional gender practices around property inheritance, responsibilities of elder care, and continuity of the family name, which privilege males in most Asian societies are largely followed even after generations of migration to the west.[9, 19, 31] Religion is implicated in son preference although the practices are neither uniform nor similar across communities. Sex selective abortions are more likely to be found among Sikh migrant communities,[12, 16] whereas Abrahamic religions (such as, Christianity and Islam), given their strictures on abortion, are more likely to have larger families until they attain the family composition of their preference.[13, 17] Although sex selective abortion is a predominant method, it is not sole form of prenatal son preference. Other methods include larger family sizes and the application of the ‘male preference stopping rule’ or the practice of stopping pregnancies when the desired male child/children have been achieved [21] and selective in-vitro fertilization practices.[32]

There is a glaring lack of comparative analyses of SRBs in Aotearoa/New Zealand despite the significant increase in the Asian migrant populations in the last 30 years. New Zealand is a demographically multi-ethnic and ‘superdiverse’ society although its politically foundations are rooted in biculturalism and *Te Tiriti o Waitangi* (The Treaty of Waitangi) signed between the Crown and Māori in 1840. According to the 2018 Census, Asian ethnicities make up around 15.1% of the total New Zealand population, a growth of approximately 10% since the 1990s.[33, 34] Asians (Chinese followed by Indians) are currently the largest ethnic minority population and comprise citizens, permanent residents, work-visa holders, refugees, and international students. Scholarship on Asian women’s lives highlights continuities in traditional gender norms.[35] In comparison to other ethnic groups, rates of induced abortion is higher among Asian women.[36, 37] In 2019, amendments to the Abortion Legislation Bill decriminalised abortion relaxing the hitherto stringent conditions for termination (see http://legislation.govt.nz/bill/government/2019/0164/latest/LMS237550.html) although Section 20F specifically notes that, “[t]his parliament opposes the performance of abortions being sought solely because of a preference for the fetus to be a of a particular sex”, requiring a review to assess evidence of any abuse of the legislation within five years.[38]

This paper, an analysis of historical and current sex ratios within and among Asian migrant populations, aims to determine the prevalence, if any, of son favouring bias in SRBs, or in other forms as part of prenatal fertility and family size decision-making.

## METHODS

### Data Sources

The Statistics New Zealand (StatsNZ) Integrated Data Infrastructure (IDI) is a secure database containing de-identified microdata on people and households, collected by government agencies and non-government organisations. Our studied utilised the Census, Ministry of Health, Department of Internal Affairs (DIA) and Ministry of Business, Innovation and Employment (MBIE) datasets.

### Historical Census Series: 1976 – 2013

Our first analysis aimed to describe the changes in ethnic sex rations over time. This analysis used Census datasets from 1976, 1981, 1986, 1991, 1996, 2001, 2006 and 2013. At the time of analysis, Census 2018 was not yet available in the IDI. From these datasets, we extracted age, sex and ethnicity data. Age was defined as age on census day. Sex was recorded as a binary variable, male or female. Ethnicity was coded according to the ethnic groups listed in the Health Information Standards Organisation’s (HISO) Ethnicity Data Protocols, the standard for collection, recording, and outputting ethnicity in the health and disability sector in New Zealand.

Of note are ethnicities that were coded for historically that are no longer coded for in more recent census. These included classifications such as West Indies, Negro, Micronesia and conglomerate ethnicities such as “Ceylonese/Singhalese/Sri Lankan”, “Syrian/Lebanese/Arab”. In the cases where there is no direct match for a historical code in the Protocols, the contemporary name of the region or name of the broad geographic area was imputed as the ethnicity code. Imputation was mostly required for MELAA and Pacific ethnicities.

Given that the historical census series is a measure of the total New Zealand population, we summarised this data as the number of males born per 100 females. To represent the range of gender ratios that are likely to arise based on random variation (which is in turn dependent on the sub-population of interest’s size), we calculated 95% confidence intervals based on binomial probability.

### Birth Register Cohort

Our birth register cohort comprised all live births between 2003 and 2018, inclusive, recorded in the DIA Births dataset. The study period was determined by the data availability of the Ministry of Health Maternity dataset. Ethnicity for the mother was obtained by linking an encrypted unique identifier associated with the birth and data in the Ministry of Health Demographics dataset.

We chose a small number of co-variates based on international literature. These were: (a) visa group, which is a proxy for generational status in New Zealand. It is assumed second and third generation migrants are likely to have resident status and those holding visas will be first generation/recent migrants. Visa status was derived from the MBIE Decisions data table; (b) parity, which was derived from the births dataset, which includes a record of the number of siblings a new born has; (c) maternal age, which was estimated based on birth year of the mother and birth year of the newborn; and (d) area level deprivation. Area level deprivation was derived using the latest notified address and linking this to an Index of Multiple Deprivation (IMD) decile. The IMD measures deprivation of a data zone in terms of employment, income, crime, housing health, education and access to essential services, and ranking each area in New Zealand from most to least deprived. The data zone is a geographic scale that is designed to capture whole neighbourhoods while preserving socioeconomic homogeneity, making it a suitable proxy for socioeconomic position. Descriptive statistics for the New Zealand birth cohort 2003-2018 are presented as counts and percentage or means with standard deviation and, medians with interquartile range. Logistic regression was used to compare odds of a male child relative to a female child and adjust for other potentially influential factors. The European/Pākeha population was used as a reference population as per other studies on SRB.

Finally, the probability of having a third child within the 2003-2018 birth cohort was examined in the context of the gender of the first two children amongst families of 3 or more children using logistic regression. Analysis was conducted for the four permutations of female and male siblings (FF, FM, MF, MM).

## RESULTS

### Historical Census series 1976-2013

The New Zealand population aged 0-5 years ranges from just over 300,000 up to 360, 000 throughout the 8 censuses between 1976 and 2013 inclusive. The Indian, Chinese, Asian and MELAA population has grown steadily as a proportion of the total over these years. The Chinese population with about 1,800 0-5-year olds in 1976 growing to 13,000 in 2013. The Indian population from 1,400 0-5-year olds in 1976 to 13,400 in 2013. The number of males for every 100 females presented by ethnicity is shown in Figure 1. The decreasing length of the 95% confidence intervals and declining variability of the male to female ratio for the Asian and MELAA populations reflects this steady growth. There is no evidence of excess female mortality, amongst the migrant population in New Zealand, in this robust data series.

**Figure 1:**
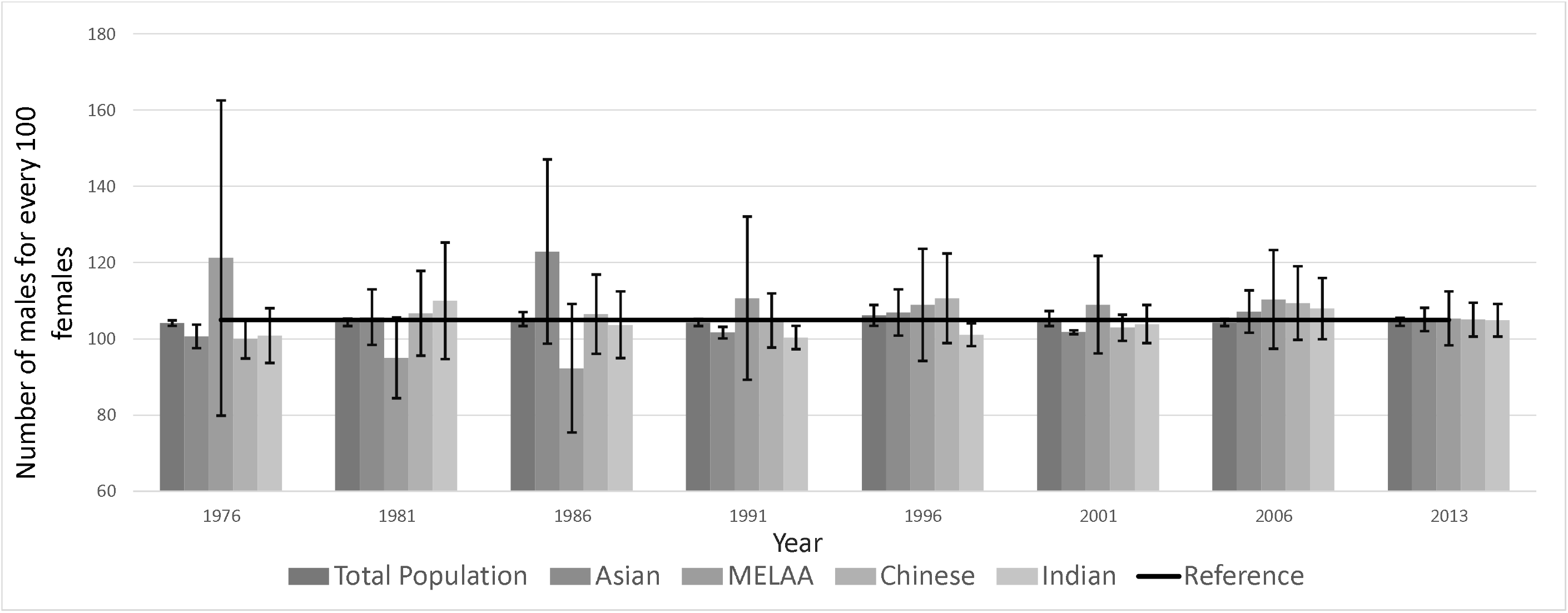
Number of males for every 100 females aged 0-5 years from NZ Census series from 1976-2013 with 95% confidence intervals and the natural expected ratio of 105 males for every 100 females.

### Birth Register Cohort 2003-2018

There were around 877,101 births in New Zealand from 2003-2018 (Table 1). Around 5% of these births were to mothers with a recent migrant & non-Pacific background. Parity is low with a median parity for the cohort of 1 and mean of 1.7 with relatively little variation amongst different ethnicities. There were no significant associations between ethnicity and odds of a male child with or without adjustment for visa group, parity, maternal age, and deprivation.

**Table 1:**
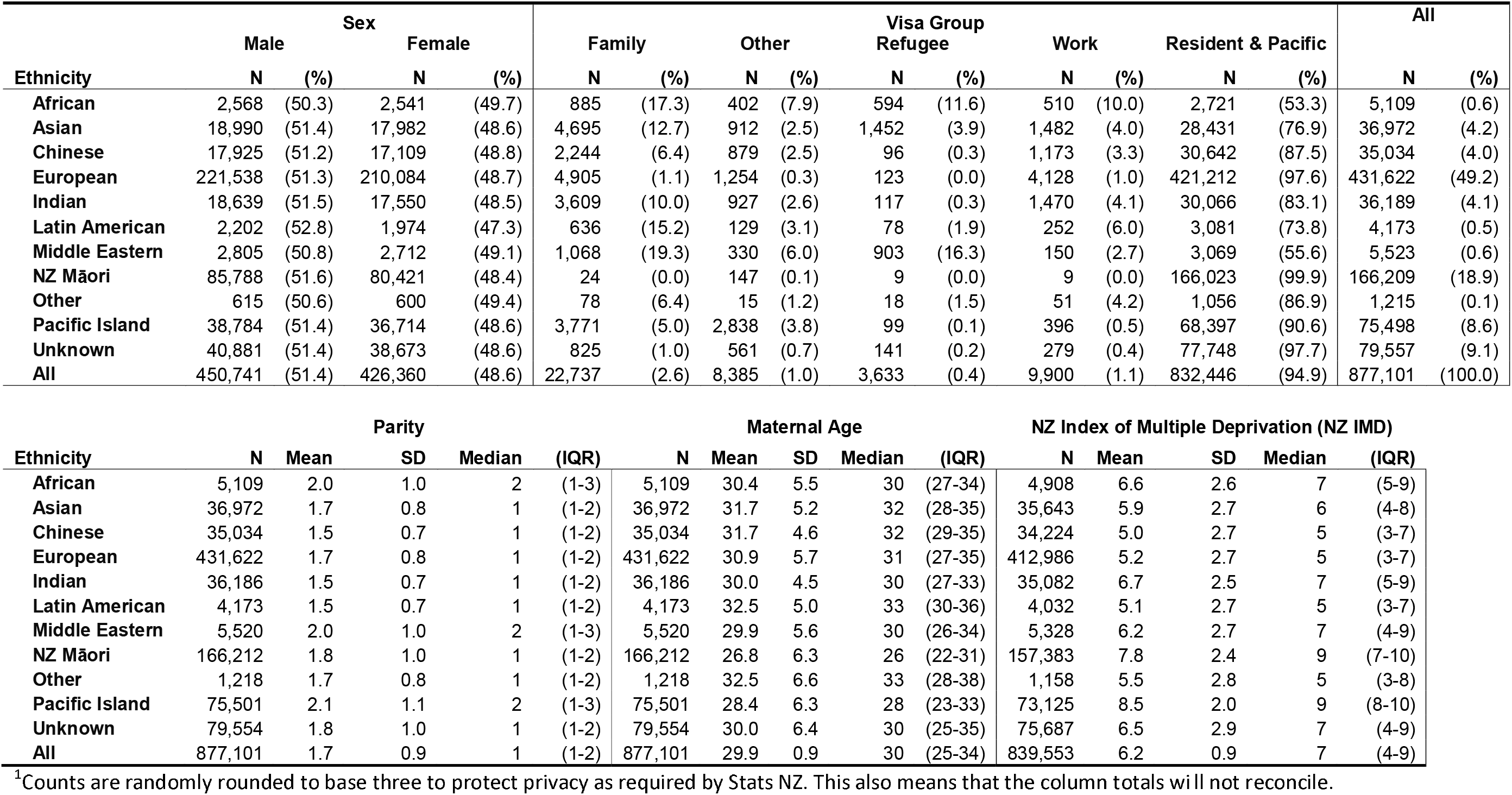
Demographic and migration characteristics of the New Zealand birth cohort from 2003 – 2018^1^.

When examining family gender composition and family size (Table 2) there was a clear preference for mixed gender regardless of ethnicity with odds of having a third child significantly lower versus two males MF vs MM, OR 0.82 (95% CI 0.80 – 0.84) and FM vs MM, OR 0.83 (95% CI 0.81-0.84). However, there was a significant interaction between family gender composition and ethnicity and therefore different ethnic groups were modelled separately. Indian families, OR 1.5 (95% CI 1.3-1.7) and Chinese families OR 1.2 (95%CI 1.1-1.4) were significantly more likely to have a third child if the first two children were female compared to both male. European families were marginally more likely to have a third child if the first two children were male, OR 0.95 (95%CI 0.93-0.98) and the significant preference for mixed gender compared to both males was maintained throughout the separate ethnic models.

**Table 2:** Percentages of families who have a third child amongst families with 2 or three children by ethnicity^1^.

|  | **African** |  |  | **Asian** |  |  | **Chinese** |  |  |
| --- | --- | --- | --- | --- | --- | --- | --- | --- | --- |
|  | **3rd child** | **(%)** | **Total** | **3rd child** | **(%)** | **Total** | **3rd child** | **(%)** | **Total** |
| **All** | 822 | (34.8) | 2364 | 4,131 | (25.4) | 16254 | 2,400 | (16.4) | 14,646 |
| **First two siblings** |  |  |  |  |  |  |  |  |  |
| **FF** | 228 | (38.2) | 597 | 1,086 | (27.9) | 3891 | 744 | (20.2) | 3,681 |
| **FM** | 186 | (33.2) | 561 | 882 | (22.8) | 3870 | 489 | (13.9) | 3,507 |
| **MF** | 189 | (33.5) | 564 | 969 | (23.8) | 4074 | 516 | (14.1) | 3,666 |
| **MM** | 219 | (34.3) | 639 | 1,197 | (27.1) | 4422 | 651 | (17.2) | 3,792 |
|  | **European** |  |  | **Indian** |  |  | **Latin American or Hispanic** |  |  |
|  | **3rd child** | **(%)** | **Total** | **3rd child** | **(%)** | **Total** | **3rd child** | **(%)** | **Total** |
| **All** | 49,500 | (25.6) | 19,3584 | 2,535 | (16.9) | 15,009 | 288 | (18.1) | 1,593 |
| **First two siblings** |  |  |  |  |  |  |  |  |  |
| **FF** | 12,732 | (27.2) | 46,743 | 867 | (22.4) | 3,876 | 69 | (19.5) | 354 |
| **FM** | 10,869 | (23.4) | 46,458 | 504 | (13.5) | 3,732 | 57 | (15.3) | 372 |
| **MF** | 10,884 | (23.1) | 47,079 | 549 | (15.3) | 3,588 | 75 | (17.4) | 432 |
| **MM** | 15,015 | (28.2) | 53,301 | 612 | (16.1) | 3,813 | 90 | (20.5) | 438 |
|  | **Middle Eastern** |  |  | **NZ Maori** |  |  | **Other** |  |  |
|  | **3rd child** | **(%)** | **Total** | **3rd child** | **(%)** | **Total** | **3rd child** | **(%)** | **Total** |
| **All** | 828 | (32.9) | 2,517 | 20,259 | (33.9) | 59,691 | 132 | (24.3) | 543 |
| **First two siblings** |  |  |  |  |  |  |  |  |  |
| **FF** | 192 | (32.7) | 588 | 5,106 | (35.9) | 14,226 | 39 | (27.7) | 141 |
| **FM** | 219 | (33.0) | 663 | 4,755 | (33.0) | 14,421 | 33 | (22.4) | 147 |
| **MF** | 204 | (34.0) | 600 | 4,704 | (32.1) | 14,667 | 24 | (22.9) | 105 |
| **MM** | 210 | (31.7) | 663 | 5,691 | (34.7) | 16,377 | 36 | (24.0) | 150 |
|  | **Pacific Island** |  |  | **Unknown** |  |  |  |  |  |
|  | **3rd child** | **(%)** | **Total** | **3rd child** | **(%)** | **Total** |  |  |  |
| **All** | 11,049 | (39.1) | 28,224 | 5,328 | (30.2) | 17,646 |  |  |  |
| **First two siblings** |  |  |  |  |  |  |  |  |  |
| **FF** | 2,703 | (40.0) | 6,756 | 1,404 | (32.8) | 4,287 |  |  |  |
| **FM** | 2,625 | (38.7) | 6,786 | 1,182 | (27.3) | 4,329 |  |  |  |
| **MF** | 2,718 | (38.2) | 7,119 | 1,200 | (27.7) | 4,335 |  |  |  |
| **MM** | 3,006 | (39.7) | 7,563 | 1,542 | (32.8) | 4,695 |  |  |  |

1Counts are randomly rounded to base three to protect privacy as required by Stats NZ. This also means that the column totals will not reconcile.

Finally, the gender ratio for parity 3 children of families with two female first born was not significantly skewed for Indian (459 males/408 females, 52.9% (Wilson 95% CI 49.6 – 56.2%) or Chinese (375 males/369 females, 50.4% (Wilson 95% CI 46.8 – 54.0%) families.

## DISCUSSION

Our study results point to marked differences in M:F ratios among New Zealand’s Asian minority and migrant population groups relative to comparator countries like Canada, the US, and Australia. Varied measures of SRBs and child sex ratios in these countries consistently demonstrated a male surplus-female deficit in Indian and Chinese populations, resulting in significant ‘missing women’ in the overall population. In contrast, child sex ratios for New Zealand Indian and Chinese ethnicity populations do not show similar male-favouring bias, and for the period under study, in the multiple data sets, i.e., Census, and Birth Data. M:F deviations from the 105:100 norm were not outside of what would be expected due to random variation. Most significantly, there was also no evidence of sex ratio skew at birth order >2/higher parities as has been seen in several other contexts. Deprivation and migration (visa status) did not have a significant influence on the gender ratio either. In all these measures, Asian (Indian and Chinese) sex ratios are unremarkable and no different to the estimates for the reference group, i.e., European/Pākeha.

The lack of evidence for excess female mortality allows us to conclude that if it occurs at all, prenatal sex-selective termination is not a widespread practice among the Asian populations of New Zealand. The absence of bias in sex ratios may be attributed to a range of factors. Prominently, New Zealand, unlike neighbouring Australia, the US or Canada, had stringent restrictions around pregnancy terminations, which may have been a deterrent to the procurement of sex-selective abortions. Also unlike in Canada and the US, where the sex of the baby may be revealed during regular ultrasound screening during pregnancy, there are stricter stipulations around such disclosures in New Zealand. Although every interaction by practitioners and their clients cannot be monitored, the evidence does not seem to suggest significant breaches in disclosures. Furthermore, ultrasounds confirming the sex of the baby are usually conducted around 18 weeks but by law abortions beyond 20 weeks are illegal. Arguably legislative restrictions are likely to affect domestic access to termination; however, there is also no evidence to suggest that Asian women travelled overseas to procure sex-selective abortions. Prohibitive costs associated with travel from New Zealand may constrain the return to home countries for this purpose but, conversely, there is no evidence to suggest that higher-income migrants have used this pathway.

More positively, acculturation of Asian migrant populations, and alignment with the more liberal gender norms in the host country, might have an influence on the reshaping of attitudes to the girl child and would be in line with results noted elsewhere.[13, 16] Broader social positioning as minority populations might have also impacted on the perception and valuation of children as assets and social capital, regardless of sex. Research has highlighted myriad ways in which 1.5 and 2^nd^ generation migrant children support families through bringing in secondary income, supporting parents through language and cultural interpretation,[39, 40] and deepening roots in the community.[39] For many first-generation migrants, children also facilitate in the process of legalising their residency status. Until 2006, any child – regardless of sex – born in New Zealand was automatically accorded citizenship. Compared to other minority groups in New Zealand, such as Māori and Pacific Island communities, or overseas such as Punjabi Sikh community in Canada, the majority of New Zealand’s Asian and ethnic community are fairly recent arrivals and are in various stages of settlement. Under these circumstances, the benefits of children, both boys and girls, outweigh any perceived cultural disadvantages.

The lack of sex-selective abortion specifically does not dismiss son preference wholly. Indeed, what has emerged from the latter part of our analysis is that desire for male children continues to be germane among Indian and Chinese couples but is practiced as extended family sizes and planned gender composition. Male preference is manifest in ‘soft’ prenatal decision-making placing it in the ethical crossroads between gender bias and sex discrimination, on the one hand, or a matter of choice and family balancing, on the other.[16, 32, 41]

### Study Limitations

Methodologically, the strengths of the study include the use of whole of population data. However this is administrative data not collected for the specific purposes of examining son preference and ‘missing women’. Therefore, there is likely to be imprecision in some of the measures, for example, visa group as a proxy of duration of family residency (generation number) in New Zealand. However, the analyses also reveal significant gaps in prenatal gender-specific data.

The analysis also used live birth sex ratios as the sole indicator of gender inequity in Asian and ethnic community. Live birth sex ratios are open to critique and needs to be triangulated with other data sets to confirm existence of sex-selective fertility practices.[32] Further work could focus on *intervals* between births, sex of *still-births*, and further disaggregation by religion, income, and region. New Zealand also currently does not capture sex-specific abortion data, especially for abortions after 14 weeks when sex is known; this too would have been a source of additional data for triangulation. Similarly, there are also comparisons to be made against the inordinate perinatal still births among the Indian population in New Zealand.[42] While as of now there is no suggestion that this gender-related, a sex-differentiated analysis would further enhance our understanding of the physiological and social implications of gender and culture during pregnancy.

Finally, the data presented here is a snapshot of time whereas the migrant community in New Zealand is continually in change. In the years to come, we are likely to have a growing proportion of migrant populations who have been born in New Zealand or have come to the country as children, and who make up the next generation of reproductive age women. There is also a rise in inter-racial/inter-ethnic marriages, all of which are likely to inform a new generation of gender norms in many of these communities.

## CONCLUSION

Our study, an exploration of M:F sex ratio imbalances in Asian immigrant populations in New Zealand, failed to reveal any skewness towards male children, or present any evidence of potential sex-selective abortion. The data, however, demonstrated extended family sizes particularly where the first two children were female.

Based on these findings, the study draws the following conclusions. The first is to underscore the ‘New Zealand anomaly’ in Asian ethnicity sex ratios disputing sex-selection and ‘missing women’ as a widespread phenomenon in the country. Secondly, we conclude that gender bias and son preference are prevalent through softer cultural practices that are not captured in sex ratio imbalances. These study findings come at an apposite moment in New Zealand’s legal history. The Abortion Legislation Bill in 2020 was enacted against the proviso that any relaxations may evoke sex selective abortion. Our results, which highlight an absence of this practice, provide a baseline for future reviews to assess the impact, if any, of the decriminalisation reforms. Instead, we argue that there should be scrutiny on the more subtle forms of son preference in the pre-natal stages including family balancing or the less explored in-vitro fertilisation practices. Equally, son preference can also be demonstrated after birth, e.g., in the opportunities available to girl children. These are areas for further research.

Most importantly, these findings suggest that culture is continually being adapted to local conditions, be it fertility regulatory frameworks or migration histories. While education programmes in the community around daughter valuation may be beneficial, son preference and its effects are likely to be ameliorated through creating social systems – for example, in education, employment, community structures, leadership and politics – that are equitable to girls and women of colour, and present the best possibilities for elimination of son preference as a cultural practice.

## Data Availability

Statistics New Zealand Disclaimer
The results in this paper are not official statistics. They have been created for research purposes from the Integrated Data Infrastructure (IDI), managed by Statistics New Zealand. The opinions, findings, recommendations, and conclusions expressed in this paper are those of the author(s), not Statistics NZ. Access to the anonymized data used in this study was provided by Statistics NZ under the security and confidentiality provisions of the Statistics Act 1975. Only people authorized by the Statistics Act 1975 are allowed to see data about a particular person, household, business, or organization, and the results in this paper have been confidentialized to protect these groups from identification and to keep their data safe. Careful consideration has been given to the privacy, security, and confidentiality issues associated with using administrative and survey data in the IDI. Further detail can be found in the Privacy impact assessment for the Integrated Data Infrastructure available from www.stats.govt.nz.

## Ethical Approval

This study was approved by the University of Auckland ethics committee on 1 July 2019 (Reference no. 023303) and formal notification that it was out of scope for HDEC review (required for use of Ministry of Health data) on 24^th^ May 2019. The paper is an output of the ‘Missing Women in New Zealand’ project funded by the Health Research Council of New Zealand (project no. 19/730).

### Statistics New Zealand Disclaimer

The results in this paper are not official statistics They have been created for research purposes from the Integrated Data Infrastructure (IDI), managed by Statistics New Zealand. The opinions, findings, recommendations, and conclusions expressed in this paper are those of the author(s), not Statistics NZ. Access to the anonymised data used in this study was provided by Statistics NZ under the security and confidentiality provisions of the Statistics Act 1975. Only people authorised by the Statistics Act 1975 are allowed to see data about a particular person, household, business, or organisation, and the results in this paper have been confidentialised to protect these groups from identification and to keep their data safe. Careful consideration has been given to the privacy, security, and confidentiality issues associated with using administrative and survey data in the IDI. Further detail can be found in the Privacy impact assessment for the Integrated Data Infrastructure available from www.stats.govt.nz.

